# Clinical and Echocardiographic Characteristics Associated with Hypertension in a Cohort of People Living with and without HIV

**DOI:** 10.64898/2026.01.12.26343986

**Authors:** David Chisompola, Sydney Mulamfu, Martin Chakulya, Emmanuel Luwaya, Phinnoty Mwansa, Benson M. Hamooya, Joreen P. Povia, Sepiso K. Masenga

**Affiliations:** Department of Cardiovascular Science and Metabolic Diseases, Livingstone Center for Prevention and Translational Science, Livingstone 10101, Zambia; HAND Research Group, School of Medicine and Health Sciences, Mulungushi University, Livingstone 10101, Zambia; Department of Public Health and Biostatistics, Livingstone Center for Prevention and Translational Science, Livingstone 10101, Zambia; Department of Health Economics, Livingstone Center for Prevention and Translational Science, Livingstone 10101, Zambia

**Author notes:** cofirst (David Chisompola, Sydney Mulamfu).

**Keywords:** Hypertension, HIV, Cardiac remodeling, Left ventricular hypertrophy, Renin-angiotensin-aldosterone system, Sub-Saharan Africa

## Abstract

**Background:** Hypertension is a major cardiovascular risk factor in sub-Saharan Africa, particularly among people living with HIV (PLWH). The contributions of cardiac structural remodeling, metabolic factors, and the renin-angiotensin-aldosterone system (RAAS) to hypertension in this population remain incompletely understood. This study aimed to provide a detailed characterization of the clinical and echocardiographic determinants of hypertension in a cohort predominantly composed of people living with HIV.

**Methods:** In this cross-sectional study, 366 adults (70.2% female, 73.5% PLWH) attending a tertiary hospital in Zambia were enrolled. Hypertension was defined as systolic/diastolic blood pressure ≥140/90 mmHg or current antihypertensive use. We collected sociodemographic, clinical, biochemical (lipid profile, RAAS markers, inflammatory cytokines), and echocardiographic data. Multivariable logistic regression models were used to identify independent predictors of hypertension, with statistical significance at p<0.05.

**Results:** Hypertension prevalence was 24.3% (n=89). Hypertensive participants were older (median age 59 vs. 45 years, p<0.0001) and had higher BMI, cholesterol, triglycerides, and left ventricular mass index (all p<0.01). In adjusted Model 1 (controlling for age, BMI, waist circumference, HIV), left ventricular septal diameter (IVSD: AOR=25.2, 95% CI: 5.7–110.2), posterior wall diameter (LVPWD: AOR=22.9, 95% CI: 4.3–121.9) and age (AOR=1.08 per year, 95% CI: 1.05–1.11) were associated with hypertension. In Model 2, adjusting for RAAS components, cardiac structural parameters and age remained significant, while RAAS biomarkers showed no independent association.

**Conclusion:** In this HIV-prevalent cohort, hypertension was independently associated with left ventricular structural remodeling and advancing age, but not with circulating RAAS components or HIV status. These findings underscore the importance of echocardiographic assessment in hypertension evaluation and suggest that non-RAAS pathways may contribute to hypertensive cardiac remodeling in this setting.

## Introduction

Hypertension remains a leading global cause of cardiovascular morbidity and mortality, with a disproportionate burden in low- and middle-income countries, particularly in sub-Saharan Africa [1]. Worldwide, the prevalence of hypertension was projected to decline modestly from 22.1% in 2015 to 20.3% (20.2 – 20.4%) by 2040 [2]. However, this decline masks a persistent and disproportionate burden in low- and middle-income countries, particularly in sub-Saharan Africa, where health systems are often strained by the dual challenges of infectious and non-communicable diseases [3]. In this region, the rising prevalence of hypertension coincides with the ongoing HIV epidemic, creating a syndemic that complicates clinical management and worsens long-term outcomes [4]. People living with HIV (PLWH) are at increased risk of hypertension due to a combination of traditional cardiovascular risk factors, chronic inflammation, immune activation, and potential metabolic effects of antiretroviral therapy (ART) [5,6].

Cardiac structural remodeling especially left ventricular hypertrophy (LVH), and increased wall thickness, is a well-established consequence of chronic hypertension and a strong predictor of adverse cardiovascular events [7,8]. However, the relative contributions of traditional risk factors, HIV-specific factors, and hormonal pathways such as the renin-angiotensin-aldosterone system (RAAS) to hypertension-associated cardiac remodeling in PLWH remain poorly characterized. While RAAS is classically associated with blood pressure regulation and fluid balance, emerging evidence suggests it also plays roles in inflammation, fibrosis, and immune modulation, pathways that may be particularly relevant in the context of HIV and chronic hypertension [9,10].

Despite these mechanistic links, few studies have comprehensively evaluated echocardiographic, metabolic, and RAAS biomarkers together in hypertensive PLWH in sub-Saharan Africa. Understanding which factors independently drive hypertension and cardiac remodeling in this population is critical for developing targeted prevention and management strategies.

This study aimed to characterize the clinical, metabolic, echocardiographic, and RAAS-related determinants of hypertension in a cohort of adults with high HIV prevalence in Zambia. Using multivariable logistic regression, we sought to identify independent predictors of hypertension and assess whether cardiac structural parameters remain significant after adjustment for traditional risk factors and circulating RAAS components.

## Materials and Methods

### Study Design and Setting

This cross-sectional study was conducted at the outpatient medical clinic of Livingstone University Teaching Hospital from October 1, 2023, to June 1, 2024. A total of 366 adults attending routine medical check-ups who provided written Informed consent were enrolled.

### Study Participants and Eligibility Criteria

Participants were eligible if they were aged ≥18 years and provided written informed consent. The cohort included individuals living with HIV (PLWH) and those without HIV (PWTH), with or without hypertension. PLWH were required to be on stable antiretroviral therapy (ART) for ≥12 months and virologically suppressed (viral load <50 copies/mL). HIV-negative status was confirmed via standard testing. Exclusion criteria comprised diabetes mellitus, pregnancy, severe renal or hepatic impairment, active opportunistic infections, or other significant comorbidities that could influence systemic inflammation or cardiovascular parameters.

### Sample Size and Sampling

A purposive (non-probability) sampling technique was employed to facilitate efficient enrollment. The sample size was calculated using the single population proportion formula. An expected prevalence of 23% [4] was assumed based on prior evidence. A 95% confidence level (Z = 1.96) and a margin of error of 5% were used. The minimum required sample size was calculated as follows:

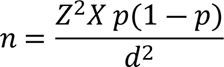

Where n represents the required sample size, p the estimated prevalence, Z the standard normal deviate corresponding to a 95% confidence level, and d the margin of error. Substitution of these values yielded a minimum sample size of 272 participants. To account for possible non-response or incomplete records, a 10% contingency was added, resulting in a final target sample size of 300 participants. To ensure robustness for subgroup analyses and account for potential data issues, we recruited 366 participants, providing ample statistical power.

### Hypertension Definition

Hypertension was defined as systolic blood pressure ≥140 mmHg and/or diastolic blood pressure ≥90 mmHg based on the average of three seated measurements taken five minutes apart [11], or current use of antihypertensive medication(s) as documented in medical records or self-reported during the study interview.

### Biochemical and Biomarker Assays

Venous blood specimens were collected, aliquoted and stored at −80°C until analysis. Inflammatory cytokines such as IL-17A, IL-6 and components of the renin-angiotensin-aldosterone system (renin, angiotensin II, aldosterone) were measured using commercial enzyme-linked immunosorbent assay (ELISA) kits according to manufacturer instructions. Lipid profiles such as total cholesterol triglycerides, and VLDL were analyzed with standard automated clinical chemistry platforms.

### Cardiac Structure Assessment

Standard transthoracic echocardiography was conducted in accordance with the American Society of Echocardiography (ASE) guidelines [13], by trained sonographers using a standardized protocol. Imaging was performed using M-mode, two-dimensional (2D), and pulsed-wave/tissue Doppler modalities. Linear cardiac dimensions, measurements included left ventricular internal diameter (LVD), interventricular septal thickness (IVSD), left ventricular posterior wall thickness (LVPWD), left ventricular mass (LVMass), left ventricular mass index (LVMI), and relative wall thickness (RWT). Left ventricular hypertrophy (LVH) was defined according to established sex-specific criteria.

### Data Collection

Sociodemographic and clinical data were collected by trained personnel using standardized instruments and entered into the Research Electronic Data Capture (REDCap) platform. Information included age, sex, marital and employment status, and medical history (HIV, hypertension, cardiovascular diseases, tuberculosis). HIV and ART details were verified through medical records.

### Statistical Analysis

Data were exported from REDCap, cleaned in Microsoft Excel, and analyzed in StatCrunch. Continuous variables are presented as median (interquartile range), and categorical variables as frequency (%). Univariate associations with hypertension were assessed using chi-square or Mann–Whitney U tests, as appropriate. Variables with p < 0.05 in univariate analysis were entered into multivariable logistic regression models to identify independent predictors. Model 1 adjusted for age, BMI, waist circumference, and HIV status. Model 2 additionally adjusted for RAAS biomarkers (renin, angiotensin II, aldosterone). Adjusted odds ratios (AOR) with 95% confidence intervals (CI) were reported. A two-tailed p < 0.05 was considered statistically significant.

### Ethical Approval

The study was approved by the Mulungushi University School of Medicine and Health Sciences Research Ethics Committee (Ref. SMHS-MU3-2023-005, July 9, 2023) and by Livingstone University Teaching Hospital management. Written informed consent was obtained from all participants prior to recruitment into the study. The study adhered to the Declaration of Helsinki and national ethical guidelines (2013). All data were anonymized, and no personal identifiable information was retained.

## Results

### Participants Characteristics

The study included a total of 366 participants, of whom 89 (24.3%) were diagnosed with hypertension. The overall cohort had a median age of 49 years (IQR: 40, 59), was predominantly female (70.2%), and the majority were living with HIV (73.5%). Key demographic and clinical characteristics stratified by hypertension status are presented in Table 1. Participants with hypertension were significantly older than those without (median [IQR]: 59 [50, 64] vs. 45 [36, 55] years; p < 0.0001) (Table 1). They also exhibited significantly higher cardiometabolic risk profiles, with greater median body mass index (27.1 vs. 23.2 kg/m²; p < 0.0001), waist circumference (92 vs. 80 cm; p < 0.0001), total cholesterol (4.8 vs. 4.2 mmol/l; p = 0.0003), triglycerides (0.97 vs. 0.75 mmol/l; p = 0.0043), and uric acid levels (383 vs. 355 µmol/l; p = 0.0493). Estimated glomerular filtration rate (eGFR) was lower in the hypertensive group (95.9 vs. 110.6 ml/min/1.73m²; p = 0.0056). Cardiac structural and functional parameters were markedly different between the groups. Hypertensive participants had significantly greater left ventricular dimensions and mass, including left ventricular internal diameter (LVD: 4.2 vs. 3.9 cm; p = 0.0006), interventricular septum thickness (IVSD: 1.2 vs. 1.0 cm; p < 0.0001), left ventricular posterior wall diameter (LVPWD: 1.2 vs. 1.0 cm; p < 0.0001), left ventricular mass index (LVMI: 101 vs. 76 g/m²; p < 0.0001), and left ventricular mass (LVMass: 187 vs. 131 g; p < 0.0001). They also had a higher relative wall thickness (RWT: 0.59 vs. 0.51; p = 0.0001). Clinical comorbidities and markers were more prevalent in the hypertensive group. The history of tuberculosis was significantly more common among hypertensive participants (51.6% vs. 23.8% among those with a TB history; p = 0.001). Hypertensive participants were also more likely to have left ventricular hypertrophy (LVH: 50.7% vs. 17.4%; p < 0.0001), hypertensive heart disease (77.8% vs. 20.1%; p < 0.0001), microalbuminuria (32.0% vs. 18.9%; p = 0.014), and to have experienced a hypertensive crisis (80.0% vs. 23.2%; p < 0.0001). Conversely, current smoking was less frequent in the hypertensive group (4.0% vs. 27.1%; p = 0.011). Notably, there were no significant differences between the groups in terms of sex, employment status, marital status, HIV status, peripheral artery disease (PAD) prevalence, ART regimen distribution, or circulating levels of inflammatory cytokines (IL-17A, IL-6) and Renin-Angiotensin-Aldosterone System (RAAS) components (Renin, Aldosterone, Angiotensin II).

**Table 1.** Participants Characteristics.

| Variable | Median (IQR)<br>/ Frequency<br>(%) | Hypertension |  | P value |
| --- | --- | --- | --- | --- |
|  |  | Yes = 89<br>(24.3) | No = 277<br>(75.7) |  |
| **Age, Years** | 49 (40, 59) | 59 (50, 64) | 45 (36, 55) | **<0.0001** |
| **Sex** |  |  |  |  |
| *Male* | 109 (29.8) | 27 (24.8) | 82 (75.2) | 0.895 |
| *Female* | 257 (70.2) | 62 (24.1) | 195 (75.9) |  |
| **Employment Status** |  |  |  |  |
| *Employed* | 121 (33.4) | 34 (28.1) | 87 (71.9) | 0.271 |
| *Unemployed* | 241 (66.6) | 55 (22.8) | 186 (77.2) |  |
| **Marital Status** |  |  |  |  |
| *Married* | 154 (42.4) | 44 (28.6) | 110 (71.4) | 0.123 |
| *Unmarried* | 209 (57.6) | 45 (21.5) | 164 (78.5) |  |
| **HIV Status** |  |  |  |  |
| *Positive* | 269 (73.5) | 65 (24.2) | 204 (75.8) | 0.909 |
| *Negative* | 97 (26.5) | 24 (24.7) | 73 (75.3) |  |
| **History of TB** |  |  |  |  |
| Yes | 31 (13.1) | 16 (51.6) | 15 (48.4) | **0.001** |
| No | 206 (86.9) | 49 (23.8) | 157 (76.2) |  |
| **PAD** |  |  |  |  |
| Yes | 58 (17.2) | 20 (34.5) | 38 (65.5) | 0.085 |
| No | 279 (82.8) | 66 (23.7) | 213 (76.3) |  |
| **ART Regimen** |  |  |  |  |
| *NNRTI+NRTI* | 4 (2.4) | 1 (25.0) | 3 (75.0) | 0.250 |
| *INSTI (TLD or TafED)* | 159 (95.8) | 44 (27.7) | 115 (72.3) |  |
| *PIs* | 3 (1.8) | 2 (66.7) | 1 (33.3) |  |
| **Smoking** |  |  |  |  |
| Yes | 25 (7.5) | 1 (4.0) | 24 (96.0) | **0.011** |
| No | 310 (92.5) | 84 (27.1) | 226 (72.9) |  |
| **Hypertension crisis on or off** |  |  |  |  |
| Yes | 10 (2.9) | 8 (80.0) | 2 (20.0) | **<0.0001** |
| No | 337 (97.1) | 78 (23.2) | 259 (76.8) |  |
| **LVH** |  |  |  |  |
| Yes | 75 (22.5) | 38 (50.7) | 37 (49.3) | **<0.0001** |
| No | 258 (77.5) | 45 (17.4) | 213 (82.6) |  |
| **Hypertensive heart Disease** |  |  |  |  |
| Yes | 27 (8.2) | 21 (77.8) | 6 (22.2) | **<0.0001** |
| No | 304 (91.8) | 61 (20.1) | 243 (79.9) |  |
| **Microalbuminuria** |  |  |  |  |
| Yes | 97 | 31 (32.0) | 66 (68.0) | **0.014** |
| No | 180 | 34 (18.9) | 146 (81.1) |  |
| **Body Mass Index Kg/m<sup>2</sup>** | 24.6 (20.8, 29.2) | 27.1 (24.0, 31.8) | 23.2 (20.2, 28.2) | **<0.0001** |
| **Duration on ART (months)** | 11 (8, 17) | 15 (10, 19) | 10 (7, 15) | **0.0008** |
| **Waist Circumference (cm)** | 83 (73, 95) | 92 (82, 103) | 80 (72, 90) | **<0.0001** |
| **IL-17A (pg/ml)** | 22.8 (18.9, 491.3) | 23 (19.9, 679.3) | 22.5 (18.8, 175.1) | 0.3592 |
| **IL-6 (pg/ml)** | 86 (0.82, 108.8) | 93.7 (0.8, 106.2) | 10.9 (0.8, 108.8) | 0.7495 |
| **Renin (pg/ml)** | 2390 (21, 3267) | 2542 (23, 3144) | 85 (20, 3292) | 0.7014 |
| **Aldosterone (pg/ml)** | 154 (84, 6167) | 198 (109, 6314) | 165 (76, 5997) | 0.2494 |
| **Angiotensin II (pg/ml)** | 1669 (41, 4876) | 1838 (52, 4764) | 1620 (39, 5203) | 0.6709 |
| **RVD (cm)** | 2.3 (2.1, 2.6) | 2.4 (2.1, 2.6) | 2.3 (2.0, 2.6) | 0.2388 |
| **LVD (cm)** | 4.0 (3.7, 4.4) | 4.2 (3.8, 4.6) | 3.9 (3.7, 4.3) | **0.0006** |
| **IVSD (cm)** | 1.1 (1.0, 1.2) | 1.2 (1.1, 1.4) | 1.0 (0.9, 1.1) | **<0.0001** |
| **LVPWD (cm)** | 1.1 (1.0, 1.2) | 1.2 (1.1, 1.4) | 1.0 (0.9, 1.1) | **<0.0001** |
| **Total Cholesterol (mmol/l)** | 4.4 (3.7, 5.1) | 4.8 (4.1, 5.4) | 4.2 (3.5, 5.0) | **0.0003** |
| **Triglycerides (mmol/l)** | 0.79 (0.55, 1.18) | 0.97 (0.63, 1.37) | 0.75 (0.53, 1.1) | **0.0043** |
| **VLDL (mmol/l)** | 0.36 (0.25, 0.54) | 0.44 (0.29, 0.62) | 0.33 (0.24, 0.49) | **0.0027** |
| **eGFR (mL/min/1.73 m<sup>2</sup>)** | 104.1 (84.1, 130.7) | 95.9 (72.7, 114.9) | 110.6 (88.2, 137.9) | **0.0056** |
| **Uric Acid (μmol/L)** | 356 (305, 416) | 383 (314, 467) | 355 (298, 403) | **0.0493** |
| **LVMI (g/m<sup>2</sup>)** | 82 (69, 100) | 101 (81, 123) | 76 (66, 91) | **<0.0001** |
| **RWT (cm)** | 0.52 (0.47, 0.60) | 0.59 (0.48, 0.68) | 0.51 (0.46, 0.57) | **0.0001** |
| **LVMass (g)** | 140 (117, 177) | 187 (137, 243) | 131 (113, 159) | **0.0001** |

Values are presented as median (interquartile range) for continuous variables and frequency (%) for categorical variables. Hypertension was defined as systolic blood pressure $\geq 140$ mmHg and/or diastolic blood pressure $\geq 90$ mmHg, or current use of antihypertensive medication. Abbreviations: IQR, interquartile range; HIV, human immunodeficiency virus; TB, tuberculosis; PAD, peripheral artery disease; ART, antiretroviral therapy; NNRTI, non-nucleoside reverse transcriptase inhibitor; NRTI, nucleoside reverse transcriptase inhibitor; INSTI, integrase strand transfer inhibitor; TLD, tenofovir/lamivudine/dolutegravir; TafED, tenofovir alafenamide/emtricitabine/dolutegravir; PIs, protease inhibitors; LVH, left ventricular hypertrophy; eGFR, estimated glomerular filtration rate; LVD, left ventricular internal diameter; IVSD, interventricular septum thickness; LVPWD, left ventricular posterior wall diameter; LVMI, left ventricular mass index; RWT, relative wall thickness; LVMass, left ventricular mass.

Figure 1 A-O, show comparisons between normotensive and hypertensive groups for: (A) age (years), (C) waist circumference (WC, cm), (D) duration on antiretroviral therapy (ART, months), (F) left ventricular diameter (LVD, cm), (G) interventricular septal diameter (IVSD, cm), (H) left ventricular posterior wall diameter in diastole (LVPWD, cm), (I) left ventricular posterior wall diameter (LVPW, cm), (J) total cholesterol (mmol/L), (K) triglycerides (mmol/L), (L) very-low-density lipoprotein (VLDL, mmol/L), (M) left ventricular mass index (LVMI, g/m²), (N) relative wall thickness (RWT), and (O) left ventricular mass (LVM, g) were statically significant. However, (B) body mass index (BMI, kg/m^2^) and (E) right ventricular diameter (RVD, cm) were not statistically significant.

**Figure 1.**
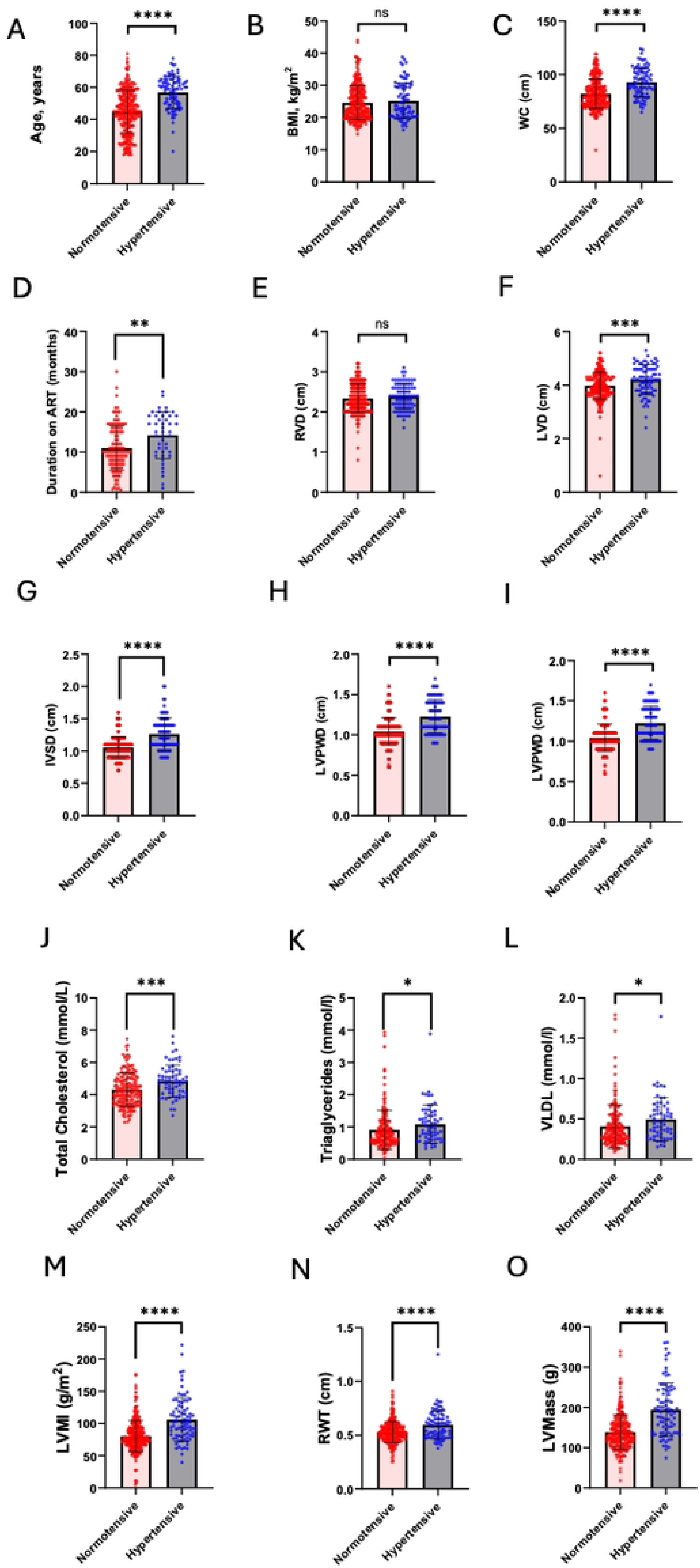
Comparison of clinical and echocardiographic parameters between Hypertensives and normotensives individuals.

### Model 1. Multivariable logistic regression analysis for factors associated with Hypertension

In model 1, a multivariable logistic regression analysis was performed to identify factors independently associated with Hypertension adjusting for age, body mass index (BMI), waist circumference, and HIV status. In the adjusted model, age remained a strong and statistically significant independent predictor, with each additional year of age associated with an 8% increase in the odds of the outcome (Adjusted Odds Ratio [AOR] = 1.08; 95% CI: 1.05, 1.11; p < 0.0001). Similarly, left ventricular mass index (LVMI) and left ventricular mass (LVMass) were independently and positively associated with the outcome (LVMI AOR = 1.02 per unit, 95% CI: 1.01, 1.03, p < 0.0001; LVMass AOR = 1.01 per unit, 95% CI: 1.00, 1.01, p < 0.0001). Several clinical cardiac parameters showed profound independent associations. The presence of IVSD and LVPWD were the strongest predictors in the adjusted model (IVSD AOR = 25.2, 95% CI: 5.7, 110.2, p < 0.0001; LVPWD AOR = 22.9, 95% CI: 4.3, 121.9, p < 0.0001). The absence of a recorded hypertensive crisis (AOR = 14.3, 95% CI: 2.27, 90.4, p = 0.005), hypertensive heart disease (AOR = 6.45, 95% CI: 2.15, 19.2, p = 0.001), and left ventricular hypertrophy (LVH) (AOR = 2.45, 95% CI: 1.29, 4.62, p = 0.006) were all independently associated with significantly higher odds of the outcome. Additionally, a history of no prior TB was associated with increased odds compared to those with a history of TB (AOR = 2.67, 95% CI: 1.05, 6.80, p = 0.039) (Table 2). Notably, the significant associations observed in univariate analysis for BMI, waist circumference, total cholesterol, and left ventricular dilatation (LVD) were attenuated and lost statistical significance after adjustment (p > 0.05). Similarly, HIV status, duration on ART, microalbuminuria, smoking status, triglycerides, VLDL, eGFR, and uric acid did not demonstrate statistically significant independent associations with the outcome in the final adjusted model. The relationship for relative wall thickness (RWT) was not statistically significant in either analysis (Table 2).

**Table 2.**
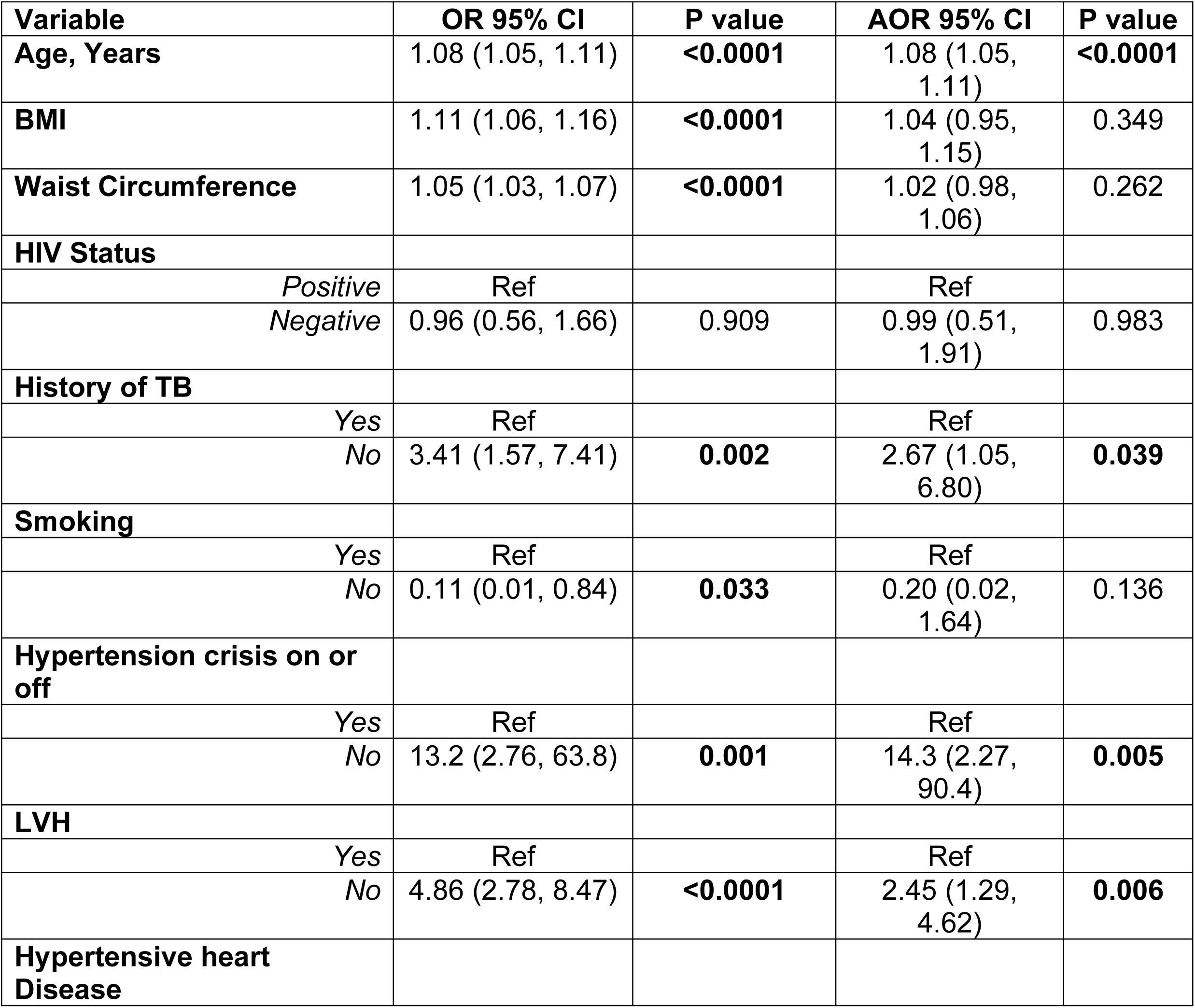
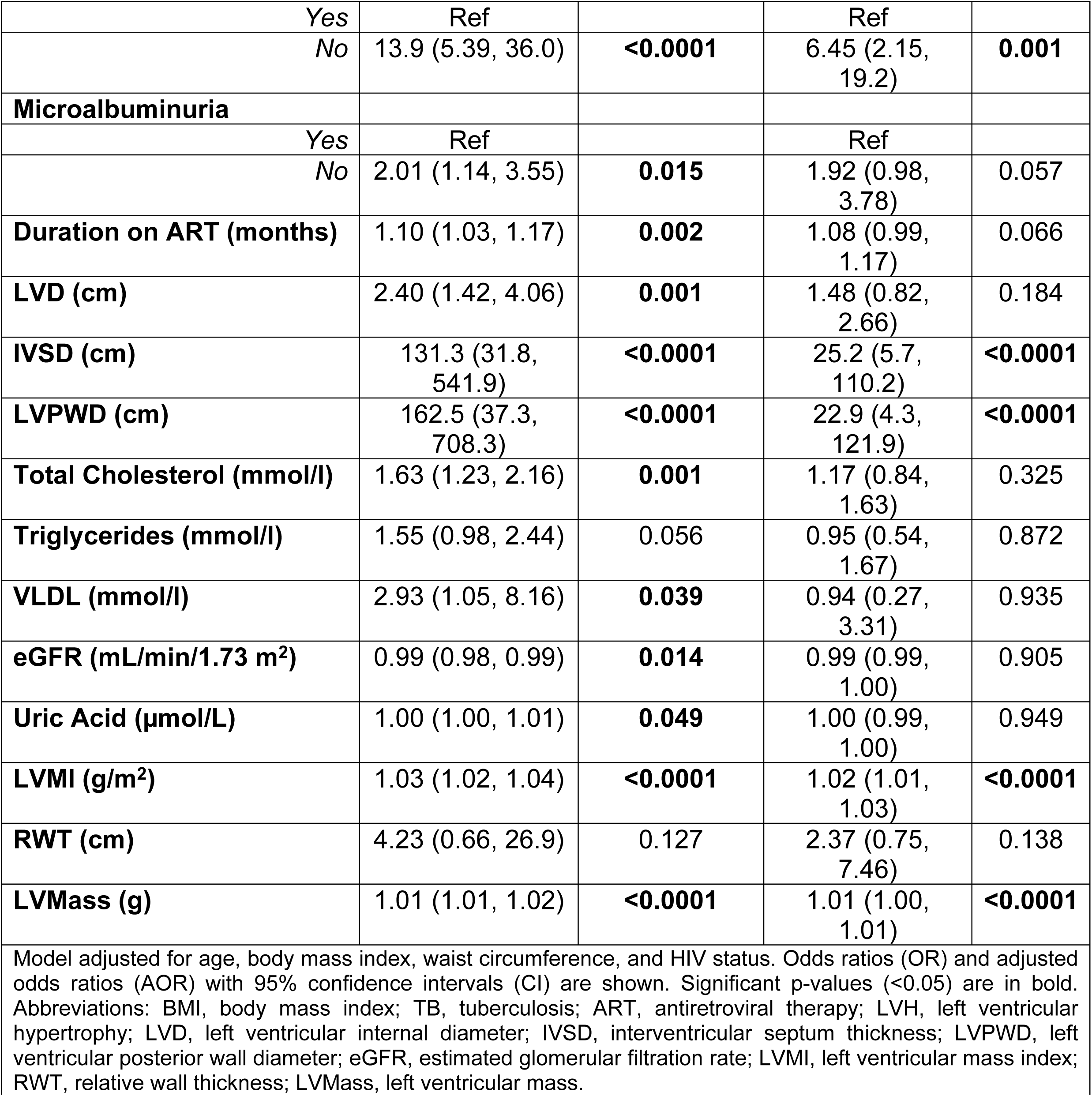
Multivariable logistic regression analysis for factors associated with Hypertension.

Figure 2 presents the final multivariable logistic regression model One. Each additional year of age was associated with an 8% increase in the odds of the outcome (AOR = 1.08; 95% CI: 1.05–1.11; p < 0.0001). Participants without a history of tuberculosis had 2.67 times higher odds compared to those with a prior TB history (AOR = 2.67; 95% CI: 1.05–6.80; p = 0.039). Individuals without a hypertensive crisis had 14.3-fold higher odds of the outcome (AOR = 14.3; 95% CI: 2.27–90.4; p = 0.005) (Figure 2). Absence of left ventricular hypertrophy (LVH) was associated with 2.45 times higher odds (AOR = 2.45; 95% CI: 1.29–4.62; p = 0.006), while absence of hypertensive heart disease was linked to 6.45-fold higher odds of the outcome (AOR = 6.45; 95% CI: 2.15–19.2; p = 0.001). Structural echocardiographic parameters showed strong independent associations. Increased interventricular septal diameter (IVSD) was associated with 25.2-fold higher odds (AOR = 25.2; 95% CI: 5.7–110.2; p < 0.0001) and increased left ventricular posterior wall diameter (LVPWD) was associated with 22.9-fold higher odds of the outcome (AOR = 22.9; 95% CI: 4.3–121.9; p < 0.0001). Measures of left ventricular mass also remained significant. Each one-unit increase in left ventricular mass index (LVMI) was associated with a 2% increase in the odds of the outcome (AOR = 1.02; 95% CI: 1.01–1.03; p < 0.0001), while each one-gram increase in left ventricular mass (LVMass) corresponded to a 1% increase in the odds (AOR = 1.01; 95% CI: 1.00–1.01; p < 0.0001). Other clinical, metabolic, and renal parameters were not independently associated with the outcome after adjustment.

**Figure 2.**
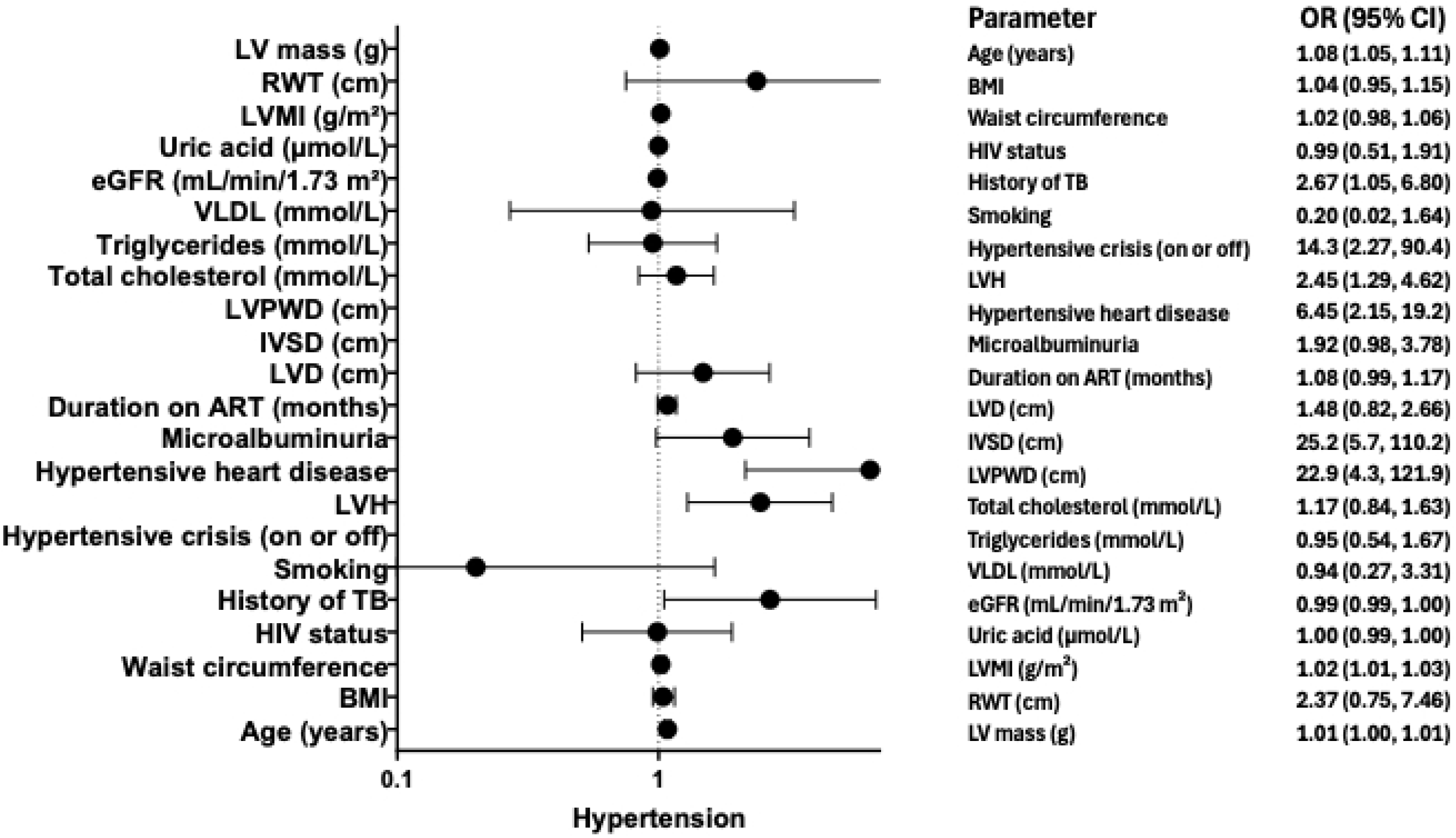
Forest plot for factors associated with Hypertension in Model One.

### Model 2. Multivariable logistic regression analysis for factors associated with Hypertension

A second multivariable model was constructed to examine the potential mediating or confounding role of the Renin-Angiotensin-Aldosterone System (RAAS). The model adjusted for age, body mass index (BMI), waist circumference, HIV status, and plasma levels of renin, angiotensin II, and aldosterone. In this RAAS-adjusted model, age remained a significant independent predictor, with each additional year associated with a 7% increase in the odds of the outcome (Adjusted Odds Ratio [AOR] = 1.07; 95% CI: 1.03, 1.11; p < 0.001). Left ventricular mass index (LVMI) and left ventricular mass (LVMass) also retained their strong, significant positive associations (LVMI AOR = 1.02, 95% CI: 1.00, 1.04, p = 0.003; LVMass AOR = 1.01, 95% CI: 1.00, 1.02, p = 0.002) (Table 3). The key echocardiographic measures of cardiac structure, IVSD) and left ventricular posterior wall diameter (LVPWD), continued to be powerful independent predictors, though the magnitude of association was attenuated compared to the first model (IVSD AOR = 16.3, 95% CI: 1.82, 146.0, p = 0.013; LVPWD AOR = 24.5, 95% CI: 1.57, 381.2, p = 0.022). The absence of hypertensive heart disease remained significantly associated with higher odds of the outcome (AOR = 4.52, 95% CI: 1.29, 15.8, p = 0.018), as did a history of no prior TB (AOR = 4.17, 95% CI: 1.07, 16.2, p = 0.039). Notably, adjustment for RAAS components led to important changes in the model. The associations for left ventricular hypertrophy (LVH) and hypertensive crisis were attenuated and lost statistical significance (p = 0.062 and p = 0.145, respectively). The direct biochemical measures of the RAAS pathway, renin, angiotensin II, and aldosterone, did not show any statistically significant independent associations with the outcome (all p > 0.2). As in the first model, the significant univariate associations for BMI, waist circumference, total cholesterol, triglycerides, VLDL, LVD, microalbuminuria, duration on ART, eGFR, and uric acid were not sustained in this multivariable analysis. HIV status and smoking status also showed no independent association. Relative wall thickness (RWT) was not significant (Table 3).

**Table 3.** Multivariable logistic regression model 2 analysis for factors associated with Hypertension.

| Variable | OR 95% CI | P value | AOR 95% CI | P value |
| --- | --- | --- | --- | --- |
| **Age, Years** | 1.08 (1.05, 1.11) | **<0.0001** | 1.07 (1.03, 1.11) | **<0.001** |
| **BMI** | 1.11 (1.06, 1.16) | **<0.0001** | 1.07 (0.93, 1.23) | 0.296 |
| **Waist Circumference** | 1.05 (1.03, 1.07) | **<0.0001** | 1.00 (0.95, 1.06) | 0.809 |
| **HIV Status** |  |  |  |  |
| Positive | Ref |  | Ref |  |
| Negative | 0.96 (0.56, 1.66) | 0.909 | 2.09 (0.23, 18.9) | 0.509 |
| **History of TB** |  |  |  |  |
| Yes | Ref |  | Ref |  |
| No | 3.41 (1.57, 7.41) | **0.002** | 4.17 (1.07, 16.2) | **0.039** |
| **Smoking** |  |  |  |  |
| Yes | Ref |  | Ref |  |
| No | 0.11 (0.01, 0.84) | **0.033** | 2.15 (0.18, 25.4) | 0.541 |
| **Hypertension crisis on or off** |  |  |  |  |
| Yes | Ref |  | Ref |  |
| No | 13.2 (2.76, 63.8) | **0.001** | 7.24 (0.50, 103.6) | 0.145 |
| **LVH** |  |  |  |  |
| Yes | Ref |  | Ref |  |
| No | 4.86 (2.78, 8.47) | **<0.0001** | 2.50 (0.95, 6.55) | 0.062 |
| **Hypertensive heart Disease** |  |  |  |  |
| Yes | Ref |  | Ref |  |
| No | 13.9 (5.39, 36.0) | **<0.0001** | 4.52 (1.29, 15.8) | **0.018** |
| **Microalbuminuria** |  |  |  |  |
| Yes | Ref |  | Ref |  |
| No | 2.01 (1.14, 3.55) | **0.015** | 1.77 (0.61, 5.10) | 0.289 |
| **Renin (pg/ml)** | 0.99 (0.99, 1.00) | 0.633 | 0.99 (0.99, 1.00) | 0.243 |
| **Aldosterone (pg/ml)** | 1.00 (0.99, 1.00) | 0.670 | 0.99 (0.99, 1.00) | 0.979 |
| **Angiotensin II (pg/ml)** | 1.00 (0.99, 1.00) | 0.889 | 1.00 (0.99, 1.00) | 0.759 |
| **Duration on ART (months)** | 1.10 (1.03, 1.17) | **0.002** | 1.12 (0.96, 1.30) | 0.143 |
| **LVD (cm)** | 2.40 (1.42, 4.06) | **0.001** | 2.37 (0.97, 5.79) | 0.058 |
| **IVSD (cm)** | 131.3 (31.8, 541.9) | **<0.0001** | 16.3 (1.82, 146.0) | **0.013** |
| **LVPWD (cm)** | 162.5 (37.3, 708.3) | **<0.0001** | 24.5 (1.57, 381.2) | **0.022** |
| **Total Cholesterol (mmol/l)** | 1.63 (1.23, 2.16) | **0.001** | 1.04 (0.67, 1.62) | 0.845 |
| **Triglycerides (mmol/l)** | 1.55 (0.98, 2.44) | 0.056 | 0.50 (0.20, 1.29) | 0.155 |
| **VLDL (mmol/l)** | 2.93 (1.05, 8.16) | **0.039** | 0.22 (0.02, 1.85) | 0.166 |
| **eGFR (mL/min/1.73 m<sup>2</sup>)** | 0.99 (0.98, 0.99) | **0.014** | 1.00 (0.99, 1.00) | 0.622 |
| **Uric Acid** | 1.00 (1.00, 1.01) | **0.049** | 0.99 (0.99, 1.00) | 0.598 |
| **LVMI (g/m<sup>2</sup>)** | 1.03 (1.02, 1.04) | **<0.0001** | 1.02 (1.00, 1.04) | **0.003** |
| **RWT (cm)** | 4.23 (0.66, 26.9) | 0.127 | 1.16 (0.19, 6.81) | 0.865 |
| **LVMass (g)** | 1.01 (1.01, 1.02) | **<0.0001** | 1.01 (1.00, 1.02) | **0.002** |

Model adjusted for age, body mass index, waist circumference, HIV status, and plasma levels of renin, angiotensin II, and aldosterone. Odds ratios (OR) and adjusted odds ratios (AOR) with 95% confidence intervals (CI) are shown. Significant p-values (<0.05) are in bold. Abbreviations: BMI, body mass index; TB, tuberculosis; ART, antiretroviral therapy; LVH, left ventricular hypertrophy; LVD, left ventricular internal diameter; IVSD, interventricular septal diameter; LVPWD, left ventricular posterior wall diameter; eGFR, estimated glomerular filtration rate; LVMI, left ventricular mass index; RWT, relative wall thickness; LVMass, left ventricular mass.

Figure 3 displays the final multivariable logistic regression model 2. with each additional year of age is associated with a 7% increase in age (adjusted OR (AOR)=1.07; 95% CI=1.03–1.11; p<0.001). History of TB was associated with 4.17 times higher odds of hypertension (AOR=4.17; 95% CI=1.07–16.2; p=0.039). Hypertensive heart Disease was associated with 4.52 times higher odds of hypertension (AOR=4.52; 95% CI=1.29– 15.8; p=0.018. Individuals with increased IVSD had 16.3-fold higher odds of the outcome (AOR = 16.3, 95% CI: 1.82–146.0; p = 0.013), while those with increased LVPWD had 24.5-fold higher odds (AOR = 24.5, 95% CI: 1.57–381.2; p = 0.022) (Figure 3).Left ventricular mass index (LVMI) and left ventricular mass (LVMass) also remained significantly and positively associated with the outcome in the adjusted model. Each one-unit increase in LVMI was associated with a 2% increase in the odds of the outcome (AOR = 1.02, 95% CI: 1.00–1.04; p = 0.003), while each one-unit increase in LVMass corresponded to a 1% increase in the odds (AOR = 1.01, 95% CI: 1.00–1.02; p = 0.002).

**Figure 3.**
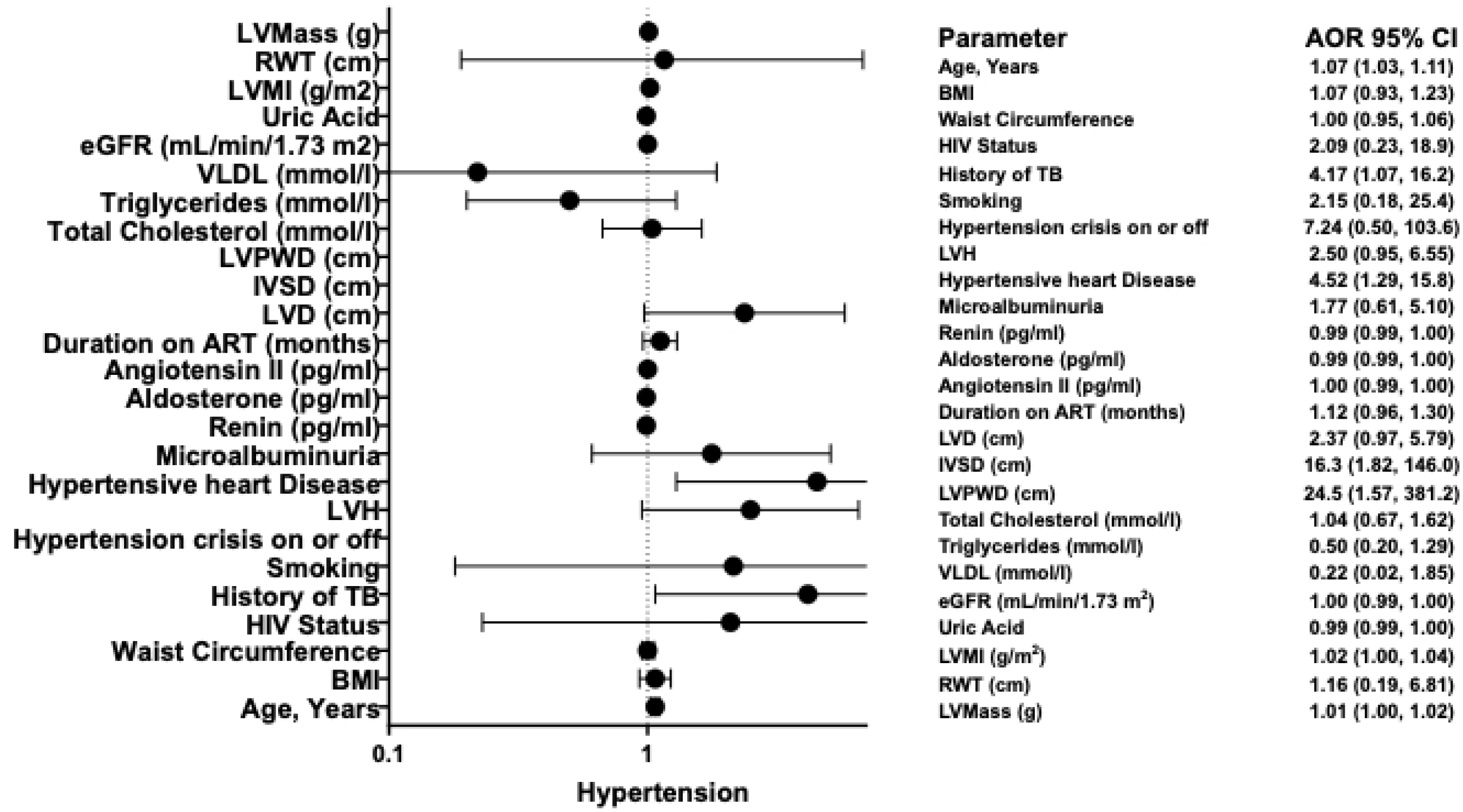
Forest plot for factors associated with Hypertension in Model Two.

## Discussion

This study aimed to provide a detailed characterization of the clinical, and echocardiographic determinants of hypertension in a cohort predominantly composed of people living with HIV. In our cohort, the overall prevalence of hypertension was 24.3% (n=89) while 24.2% (n=65) among people living with HIV. Our multivariable analysis (Model 1) identified advancing age and cardiac structural remodeling, specifically left ventricular hypertrophy (LVH) and increased wall thickness (IVSD and LVPWD), as strong independent predictors of hypertension. In contrast, HIV status, smoking status, and relative wall thickness (RWT) were not significantly associated with hypertension in the adjusted model. Further adjustment for the renin-angiotensin-aldosterone system (RAAS) in Model 2 attenuated certain clinical associations: hypertensive heart disease and hypertensive crisis lost statistical significance, while RWT remained non-significant. These findings suggest that while cardiac remodeling and age are robust predictors of hypertension in this population, the contribution of RAAS and certain clinical comorbidities may be mediated through or confounded by these structural changes.

The prevalence of hypertension observed in our study (24.3%) is comparable to rates reported in similar populations in India (23.8%) [12], Nigeria (24.9%) [13], and Kenya (25%) [3]. However, our estimate is higher than the (18.4%) and (22.5%) previously reported in Zambia [4,14] and lower than the 37.5% documented in Ethiopia. Although hypertension did not differ significantly between PLWH and PWTH in our cohort, the observed regional variability in prevalence may be attributable to differences in sample demographics, including variations in age distribution, duration of HIV infection, and exposure to specific ART regimens across studies. Additionally, environmental, genetic and lifestyle factors such as smoking prevalence, and methodological differences in hypertension ascertainment could further explain the observed variability [15].

In both multivariable models, age remained a powerful independent predictor of hypertension, with each additional year increasing the odds by approximately 7–8%. This aligns with global epidemiological data showing a rising prevalence of hypertension with aging due to vascular stiffening, endothelial dysfunction, and cumulative exposure to risk factors [16,17]. Cardiac structural parameters, especially IVSD and LVPWD, emerged as the strongest echocardiographic predictors of hypertension. These findings underscore the central role of left ventricular remodeling in hypertensive disease, even after adjustment for body mass index, HIV status, and RAAS components. Similarly, increased LVMI and LVMass were consistently and independently associated with hypertension, reinforcing the clinical relevance of echocardiography in hypertension assessment.

A history of no prior tuberculosis was unexpectedly associated with higher odds of hypertension in adjusted analyses. Although this observation aligns with findings from previous studies [18,19], it remains counterintuitive and warrants further investigation. it may reflect differences in healthcare access, chronic inflammation, or residual confounding. Conversely, hypertensive heart disease and hypertensive crisis (in Model 1) were strongly associated with hypertension, confirming the clinical relevance of these disease manifestations. The loss of significance for hypertensive crisis in Model 2 suggests possible interaction with RAAS activity, though RAAS components themselves (renin, angiotensin II, aldosterone) were not independently significant. Beyond its cardiovascular effects, the renin-angiotensin-aldosterone system also significantly contributes to inflammation, fibrosis, immune modulation, tumorigenesis, and tissue-specific disease processes [20]. However, it is not clear on mechanistic processes underlying these effects.

HIV status and duration on ART were not independently associated with hypertension in multivariable models, indicating that HIV itself may not be a primary driver of hypertension in this cohort once cardiac structure and age are accounted for. This is consistent with some contemporary studies suggesting that traditional cardiovascular risk factors, rather than HIV-specific factors dominate hypertension risk in well-controlled HIV populations [21].

The inclusion of RAAS components in Model 2 did not meaningfully alter the associations of cardiac structure with hypertension, nor did renin, angiotensin II, or aldosterone show independent effects. This suggests that in this cohort, hypertension-related cardiac remodeling may occur through RAAS-independent pathways, or that local tissue RAAS activity (not captured by plasma levels) plays a more important role. This finding highlights the complexity of hypertension pathophysiology and the potential limitation of circulating RAAS biomarkers in reflecting tissue-level activity.

Our findings have important implications for hypertension management in settings with high HIV prevalence. First, the strong independent association between hypertension and echocardiographic markers of left ventricular remodeling, particularly interventricular septal and posterior wall thickness, supports the integration of routine echocardiography into hypertension risk stratification in similar populations. Early identification of cardiac structural changes could guide more aggressive blood pressure management and closer monitoring for heart failure risk. Second, the lack of association between hypertension and circulating RAAS components suggests that RAAS-independent pathways may be prominent in hypertensive cardiac remodeling in this group, highlighting the potential need for tailored therapeutic strategies beyond RAAS inhibition. Finally, since HIV status itself did not independently predict hypertension, cardiovascular risk prevention in PLWH should emphasize control of traditional risk factors, such as obesity and dyslipidemia, alongside HIV-specific care. Future interventions could prioritize blood pressure control in older adults and those with evidence of cardiac remodeling, regardless of HIV serostatus.

Several limitations should be considered. First, the cross-sectional design precludes causal inference. Second, the sample size, though adequate for many analyses, may have limited power to detect modest associations, especially in subgroup analyses. Third, plasma RAAS components were measured at a single time point and may not reflect chronic activity. Fourth, residual confounding by unmeasured factors such as dietary salt intake, physical activity, and chronic kidney disease is possible. Finally, the generalizability may be limited to similar HIV-prevalent settings in sub-Saharan Africa.

### Conclusion

This study identifies left ventricular structural remodeling, particularly increased septal and posterior wall thickness and age as the most robust independent predictors of hypertension in a cohort with high HIV prevalence. Metabolic factors such as obesity and dyslipidemia appear to exert their influence largely through these cardiac changes. The lack of independent association with circulating RAAS components suggests that non-RAAS pathways may be important in hypertension-related cardiac remodeling in this population. These findings support the integration of echocardiography into hypertension risk stratification and management programs in similar settings and highlight the need for longitudinal studies to clarify causal pathways and inform targeted interventions.

## Availability of data and materials

The data are not publicly available due to privacy or ethical restrictions. The data will be made available upon request to qualified researchers, subject to a formal data sharing agreement and approval by our Institutional Review Board, which ensures data will be handled in accordance with the ethical restrictions under which they were collected. To access the data please contact the Mulungushi University School of Medicine and health Sciences Review board, Akapelwa street, Livingstone, Zambia 10101; Phone +260 967758554;

## Competing interests

The authors declare no competing interest exist.

## Funding

This work was supported by the Fogarty International Center and National Institute of Diabetes and Digestive and Kidney Diseases of the National Institutes of Health grants R21TW012635 (SKM), and the American Heart Association Award Number 24IVPHA1297559 (SKM).

## Supporting information

S1. Strobe Checklist

## Notes

### Competing Interest Statement

The authors have declared no competing interest.

### Funding Statement

Yes

